# Equity and coverage in RMNCH health interventions by ethnicity, 2004- 2018: lessons learned from integral policies in Ecuador

**DOI:** 10.1101/2024.10.04.24314912

**Authors:** Paulina Ríos-Quituizaca, Leonardo Ferreira, Jesus Endara-Mina, Nancy Armenta

## Abstract

**Introduction:** In Latin America, ethnic disparities rooted in the colonial legacy have persisted. There is limited scientific evidence critically analyzing the temporal changes in ethnic inequalities in reproductive, maternal, neonatal, and child health (RMNCH), and the potential impact of policies on the progress of reducing these gaps for Indigenous peoples. After the 2000 crisis, Ecuador had the region’s largest ethnic disparities in intervention coverage and social determinants due to structural racism. Inclusion policies improved starting in 2008. The main study aim was to analyze the coverage and ethnic inequalities of six RMNCH health interventions, the evolution of social determinants, and the potential impact of policies and strategies over 14 years.

**Methods:** using a mixed method design, we analyze three nationally representative surveys (2004, 2012, and 2018) to compare the evolution of social determinants; and the coverage and inequalities of six RMNCH health interventions, stratified by ethnicity (women and child Indigenous, Afro-Ecuadorians, and reference group); and we estimated absolute inequality measures and adjusted coverage ratios using Poisson regression models. Through a literature review and temporal graphs, we analyzed plans, policies, and strategies in health, education, and ethnic inclusion during the same period to estimate their potential impact.

**Results:** By 2018, the population self-identifying as Indigenous had doubled the percentage of people in the 4th and 5th wealth quintiles (from 10% to 20%) and increased secondary education attainment from 25% to 45% compared to 2004, though these changes were less evident in rural areas. Between 2004 and 2018, prenatal care coverage, institutional delivery care, and births attended by professionals increased from 27% to 75% among the Indigenous population. Although the coverage ratios show that ethnic gaps persist, inequalities progressively reduced during this period. These reductions coincided with efforts of social participation and organization that led to the constitutional recognition of pluractionality, combined with increased social investment in education and health in rural areas, and the development and implementation of policies and strategies that promoted intercultural practices in health. However, there is a noted lack of monitoring processes and impact assessment of these strategies.

**Conclusions:** The reduction of ethnic inequalities in RMNCH in Ecuador could be explained by inclusive policies and programs implemented in recent years, social organization and participation, and the involvement of actors and leaders in the implementation of these. Temporal studies based on routine surveys allow for the observation of changes and analysis of the potential impact of policies and strategies. Ecuador exemplifies actions that may have contributed to the reduction of inequalities, which could serve as a reference for other countries seeking to improve the health of Indigenous peoples. These observations also provide a pre-pandemic image, offering insights prior to the potential effects of COVID-19 and five years before assessing the outcomes of the 2030 Agenda for Sustainable Development.

## Background

In Latin America, ethnic disparities rooted in the colonial legacy have been persistent, as evidenced by unequal coverage in health interventions. Despite ethnic affiliation being one of the primary social determinants of health, there is limited available information analyzing ethnic gaps in maternal and child health (1–3).

To enhance the analytical capacity of countries making progress towards the Sustainable Development Goals (SDGs), national legislation should be implemented that aligns with fundamental principles (8) In-depth analysis of countries is also recommended to understand the factors driving success in reducing ethnic disparities (9). In the Latin American context, few countries have managed to identify both an increase in coverage and a reduction in inequality gaps over time (4). Several studies have shown that the reduction of inequalities is associated with increased investment in education, more equitable social spending, and the implementation of social policies that support vulnerable population subgroups (5–7). Our previous analysis showed that Ecuador significantly increased RMNCH intervention coverage and reduced ethnic inequalities between 2004 and 2012. Despite this, RMNCH coverage remained lower among Indigenous people until 2012. In 2008, Ecuador reinforced the guarantee of rights for indigenous peoples with the promulgation of the Constitution (8).

Studies that have expanded the temporal window of analysis for health intervention coverage among different ethnic groups, along with an examination of implemented policies and the inclusion of social determinant conditions, provide a clearer understanding of the laws, policies, or programs that may have influenced the observed temporal changes (7,9). With just over five years left to assess the outcomes of the 2030 Agenda for Sustainable Development and in the aftermath of the global COVID-19 pandemic, the most recent round of the 2018 ENSANUT survey in Ecuador, offers an opportunity to evaluate progress in reducing inequalities and to understand the situation before the pandemic. This study has an objective to compare sociodemographic characteristics by ethnicity, between 2004 and 2018; to analyze the coverage and inequalities by ethnic group in six RMNCH health interventions, and finally to analyze the policies implemented throughout this period, both in the rights of Indigenous peoples as well as policies and programs in the health sector.

## Methods

We conducted a mixed-methods case study, that analyzes data from three nationally representative health surveys conducted between 2004 and 2018, along with a literature review of the main equality policies and strategies targeting the ethnic population during the study period.

### Study design, data sources and participants

This cross-sectional study uses the Reproductive Health Survey (RHS) 2004 and the National Health and Nutrition Survey (ENSANUT) 2012 and 2018. Each survey used multistage cluster sampling to obtain nationally representative data. The datasets analyzed during this study are publicly available: the RHS 2004 in the repository of the World Bank database and the ENSANUT 2012 and 2018 in the National Institute of Statistics and Censuses of Ecuador (INEC). Ethical approval was obtained from the national agencies responsible for each survey. All analyses relied on publicly available, anonymized databases; ethical approval was not required. We followed STROBE guidelines (see Supplementary Table 1).

By the established methodology for every official survey report, one person per age group was randomly selected from each household: a woman of childbearing age (15 to 49 years old) and a child under five years. We used the population size estimates of women aged 15– 49 years and children aged 12–23 months, for whom data were available in the included surveys

#### Ethnicity

We examined inequalities by ethnicity according to the “self-identification” criterion, which was deemed the most appropriate instrument to assess the magnitude of the Indigenous and Afro-descendant population. We used three broad categories in the analyses: Indigenous, afro-descendent, and reference group (mixed ancestry (“mestizo”) or European descent (“blanco”). Our study excluded the group who self-identified as “montubios” to keep the comparability because it was not recognized as a self-identification category in the 2004 survey.

#### Coverage indicators

We assessed six essential coverage indicators covering the continuum of care for Reproductive, Maternal, Newborn, and Child Health (RMNCH): use of modern contraceptives, Antenatal care (4+ visits), Skilled birth attendance, Institutional delivery, Early initiation of breastfeeding, and Full immunization. We used standardized indicator definition criteria to ensure comparability throughout the surveys. The coverages for each indicator were defined as the percentage of women or children who receive a specific intervention among those who need it. All indicator definitions are available in Supplementary Table 2.

#### Statistical analysis

We described the national coverage prevalence according to ethnicity for each health indicator and stratified by area of residence (urban-rural), women’s education, and wealth quintiles. To measure the absolute inequality by ethnicity, we used the “mean difference from the best-performing subgroup,” where the larger values indicate higher levels of inequality, and zero indicates the absence of inequality. Moreover, we visually represented the coverage level and gaps between ethnic groups through an Equiplot.

We assessed relative ethnic inequality by calculating adjusted coverage ratios (CR) with Poisson regression models for outcomes defined as binary variables. We used the adjusted models based on household wealth, women’s education, or urban-rural residence. The robust variance option for Poisson regression ensures that the assumptions behind the regression model are not violated. All analyses considered the survey design and were conducted using Stata 17.

### Desk review

The team conducted a rapid review of existing literature on the strategies, laws, and political changes affecting the indigenous peoples of Ecuador (including indigenous and Afro-Ecuadorian populations), along with an assessment of policies and programs related to Maternal, Neonatal, Child, and Adolescent Health (RMNCH). The review took place between October to November 2023. It focused on publications and documents issued between 2000 and 2024 to actuality. The year 2000 was chosen as a starting point due to the first analyzed national health survey (ENDEMAIN 2004). Databases such as MEDLINE (via PubMed) and BIREME (via Lilacs) were used. Additionally, official websites of the Ecuadorian government, and reports from international organizations, such as the Pan-American Health Organization (PAHO), or United Nations Population Fund (UNFPA) were incorporated. Google Scholar was used to ensure the inclusion of gray literature from other academic sources.

For the initial search focused on laws, plans, projects, and policies related to Ecuador’s Indigenous peoples, the following terms were used: “laws,” “plans,” “projects,” and “policies,” combined with “peoples and nationalities,” “plurinational,” and “Ecuador.” In the second search, which focused on reproductive and maternal-child health, the terms used were: “laws,” “plans,” “projects,” “policies,” combined with “reproductive health,” “maternal-child health,” and “Ecuador.” Subsequently, abstracts were scrutinized applying predefined inclusion and exclusion criteria aligned with the research objectives. A qualitative content analysis approach was used to identify and categorize the main strategies, laws, and political changes, as well as RMNCH policies and programs implemented in Ecuador between 2000 to 2018.

## Results

Without missing data, the tree surveys allowed us to analyze data from 64,726 women and 34,100 children aged 12–23 months. Between 2004 and 2012, the distribution of the population self-identifying as Indigenous remained quite similar, but with a higher proportion of illiteracy compared to other ethnic groups. However, from 2012 to 2018, the secondary education level nearly doubled (from approximately 25% to 48%), and the illiteracy rate decreased from 18% to 4%. Similarly, the distribution of this population by wealth quintiles remained unchanged between 2004 and 2012, but by 2018, the percentages in quintiles 4 and 5 had doubled (from 10% to 20% in Q1 and Q2). The proportion of the population living in urban areas doubled between 2004 and 2012 (from 18% to 36%), but from 2012 to 2018, the increase was only 10%. The distribution of the weighted number of women by ethnic group and stratified is seen in Supplemental Table 3.

**Figure 1.**
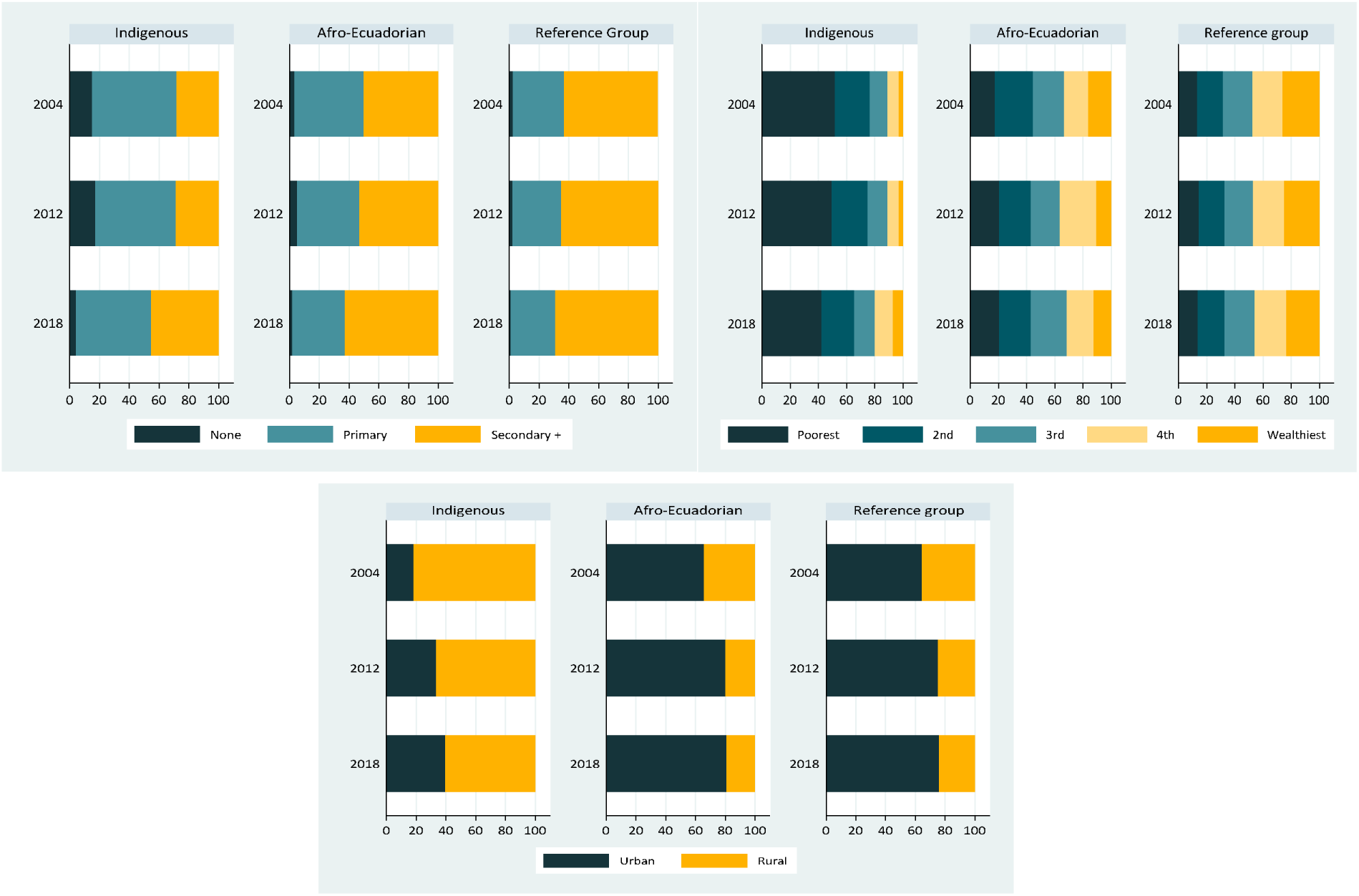
Socio-demographic characteristics of women by ethnicity (2004-2018)

In the Afro-Ecuadorian population, similar to the reference group, no significant changes in education levels were observed between 2004 and 2012. However, between 2012 and 2018, there was a noticeable reduction in illiteracy and an increase in the proportion of the population with secondary education or higher. Unlike the Indigenous population, the percentages in wealth quintiles 4 and 5 slightly decreased (from 38% to 34%). Migration to urban areas increased between 2004 and 2012 and remained stable through 2018.

Compared to other ethnic groups, despite the observed improvements, by 2018, the Indigenous population continued to predominantly reside in rural areas, had the lowest percentage of individuals with secondary education or higher, and the proportion in the lowest poverty quintile (Q1) was four times greater compared to the lowest poverty quintile in the reference group. In contrast, the Afro-Ecuadorian population is predominantly urban, with a level of secondary education or higher like the reference population, and the proportion in the highest poverty quintile is twice that of the highest poverty quintile in the reference group.

**Table 1.** Coverage and magnitude of inequalities by ethnic group, in six interventions in Ecuador from 2004 to 2018.

| RMNCH Intervention | Year | Overall |  | Indigenous group |  | Afroecuadorian group |  | Reference group |  | Difference Afroecuadorian - reference |  | Difference Indigenous-reference |  | Mean difference from best |  |
| --- | --- | --- | --- | --- | --- | --- | --- | --- | --- | --- | --- | --- | --- | --- | --- |
|  |  | % | (CI 95%) | % | (CI 95%) | % | (CI 95%) | % | (CI 95%) | % | (CI 95%) | p.p. | (CI 95%) | p.p. | (CI 95%) |
| Antenatal care 4+ visits | 2004 | 69,5 | 66,4 ; 72,4 | 27,3 | 22,4 ; 32,7 | 71,0 | 63,3 ; 77,7 | 74,0 | 50,5 ; 65,8 | -3,0 | -11,5 ; 5,5 | -46,7 | -55,5 ; -38,0 | 16,6 | 12,1 ; 21,1 |
|  | 2012 | 86,8 | 84,9 ; 88,5 | 58,3 | 50,5 ; 65,8 | 85,6 | 77,8 ; 91,0 | 89,4 | 87,6 ; 90,9 | -3,7 | -4,0 ; -3,4 | -31,0 | -31,4 ; -30,7 | 11,6 | 11,4 ; 11,8 |
|  | 2018 | 89,3 | 88,5 ; 90,1 | 74,5 | 71,3 ; 83,7 | 88,9 | 83,7 ; 92,6 | 90,9 | 90,0 ; 91,7 | -2,0 | -4,3 ; 0,3 | -16,3 | -18,6 ; -14,1 | 6,1 | 4,2 ; 8,0 |
| Skilled birth attendance | 2004 | 72,0 | 68,6 ; 75,1 | 27,6 | 21,7 ; 34,5 | 67,9 | 56,6 ; 77,4 | 78,0 | 74,9 ; 80,8 | -10,1 | -17,3 ; -2,8 | -50,3 | -57,2 ; -43,5 | 20,1 | 16,6 ; 23,7 |
|  | 2012 | 89,5 | 88,1 ; 90,7 | 53,9 | 45,6 ; 62,0 | 87,6 | 81,9 ; 91,6 | 93,0 | 91,5 ; 94,3 | -5,4 | -5,7 ; -5,2 | -39,1 | -39,5 ; -38,8 | 14,9 | 14,6 ; 15,1 |
|  | 2018 | 96,3 | 95,7 ; 96,8 | 77,7 | 73,9 ; 81,0 | 94,2 | 88,7 ; 97,1 | 98,4 | 98,0 ; 98,7 | -4,2 | -5,8 ; -2,7 | -20,7 | -22,7 ; -18,8 | 8,3 | 6,9 ; 9,7 |
| Institutional delivery | 2004 | 73,6 | 70,3 ; 76,7 | 27,8 | 22,0 ; 34,5 | 69,7 | 56,1 ; 80,5 | 74,9 | 70,9 ; 78,5 | -5,2 | -10,5 ; 0,1 | -47,1 | -52,2 ; -42,0 | 17,4 | 14,7 ; 20,1 |
|  | 2012 | 90,2 | 88,0 ; 92,1 | 55,0 | 47,0 ; 62,6 | 87,9 | 82,2 ; 91,9 | 94,0 | 92,4 ; 95,3 | -6,2 | -6,4 ; -5,9 | -39,1 | -39,4 ; -38,7 | 15,1 | 14,9 ; 15,3 |
|  | 2018 | 95,6 | 94,9 ; 96,1 | 76,6 | 72,8 ; 87,9 | 93,2 | 87,9 ; 96,3 | 97,7 | 97,2 ; 98,1 | -4,5 | -6,2 ; -2,8 | -21,1 | -23,1 ; -19,2 | 8,5 | 7,1 ; 10,0 |
| Use of modern contraceptive | 2004 | 58,4 | 56,4 ; 60,4 | 24,3 | 19,5 ; 29,8 | 57,4 | 49,9 ; 64,6 | 61,3 | 59,3 ; 63,2 | -3,8 | -12,8 ; 5,1 | -37,0 | -44,8 ; -29,3 | 13,6 | 9,6 ; 17,6 |
|  | 2012 | 70,9 | 69,2 ; 72,5 | 52,9 | 48,9 ; 56,8 | 67,0 | 58,3 ; 74,7 | 72,0 | 70,2 ; 73,6 | -5,0 | -5,3 ; -4,6 | -19,1 | -19,4 ; -18,7 | 8,0 | 7,8 ; 8,2 |
|  | 2018 | 72,4 | 90,1 ; 73,4 | 58,8 | 55,7 ; 61,9 | 72,9 | 68,2 ; 77,1 | 73,1 | 71,9 ; 74,3 | -0,3 | -3,4 ; 2,8 | -14,3 | -16,7 ; -11,9 | 4,9 | 2,6 ; 7,1 |
| Full immunization | 2004 | 53,0 | 48,0 ; 58,0 | 37,3 | 23,3 ; 53,7 | 47,1 | 26,2 ; 69,0 | 55,6 | 49,9 ; 61,2 | -8,5 | -24,7 ; 7,7 | -18,3 | -29,5 ; -7,1 | 8,9 | 2,3 ; 15,6 |
|  | 2012 | 66,6 | 62,9 ; 70,1 | 61,7 | 50,4 ; 71,9 | 60,7 | 46,6 ; 73,2 | 68,1 | 64,2 ; 71,8 | -7,4 | -8,3 ; -6,5 | -6,4 | -7,1 ; -5,7 | 4,6 | 4,0 ; 5,2 |
|  | 2018 | 70,6 | 68,2 ; 72,9 | 62,8 | 54,6 ; 70,3 | 69,2 | 60,9 ; 76,4 | 71,5 | 68,8 ; 74,0 | -2,3 | -9,5 ; 5,0 | -8,7 | -13,6 ; -3,8 | 3,7 | -1,1 ; 8,4 |
| Early initiation of breastfeeding | 2004 | 29,1 | 25,9 ; 32,5 | 36,4 | 30,2 ; 43,2 | 17,9 | 10,0 ; 30,0 | 27,9 | 24,7 ; 31,3 | -9,9 | -28,8 ; 8,9 | 8,6 | 4,2 ; 13,0 | 9,0 | 5,5 ; 12,5 |
|  | 2012 | 52,6 | 49,7 ; 55,5 | 69,6 | 62,7 ; 75,8 | 62,4 | 52,8 ; 71,2 | 51,2 | 48,2 ; 54,2 | 11,3 | 10,6 ; 12,0 | 18,4 | 18,0 ; 18,9 | 8,5 | 8,3 ; 8,8 |
|  | 2018 | 71,1 | 69,3 ; 72,8 | 77,0 | 72,2 ; 81,1 | 80,2 | 74,0 ; 85,3 | 69,9 | 67,9 ; 71,8 | 10,3 | 5,9 ; 14,8 | 7,1 | 3,8 ; 10,3 | 5,8 | 2,2 ; 9,4 |

**Table 2.**
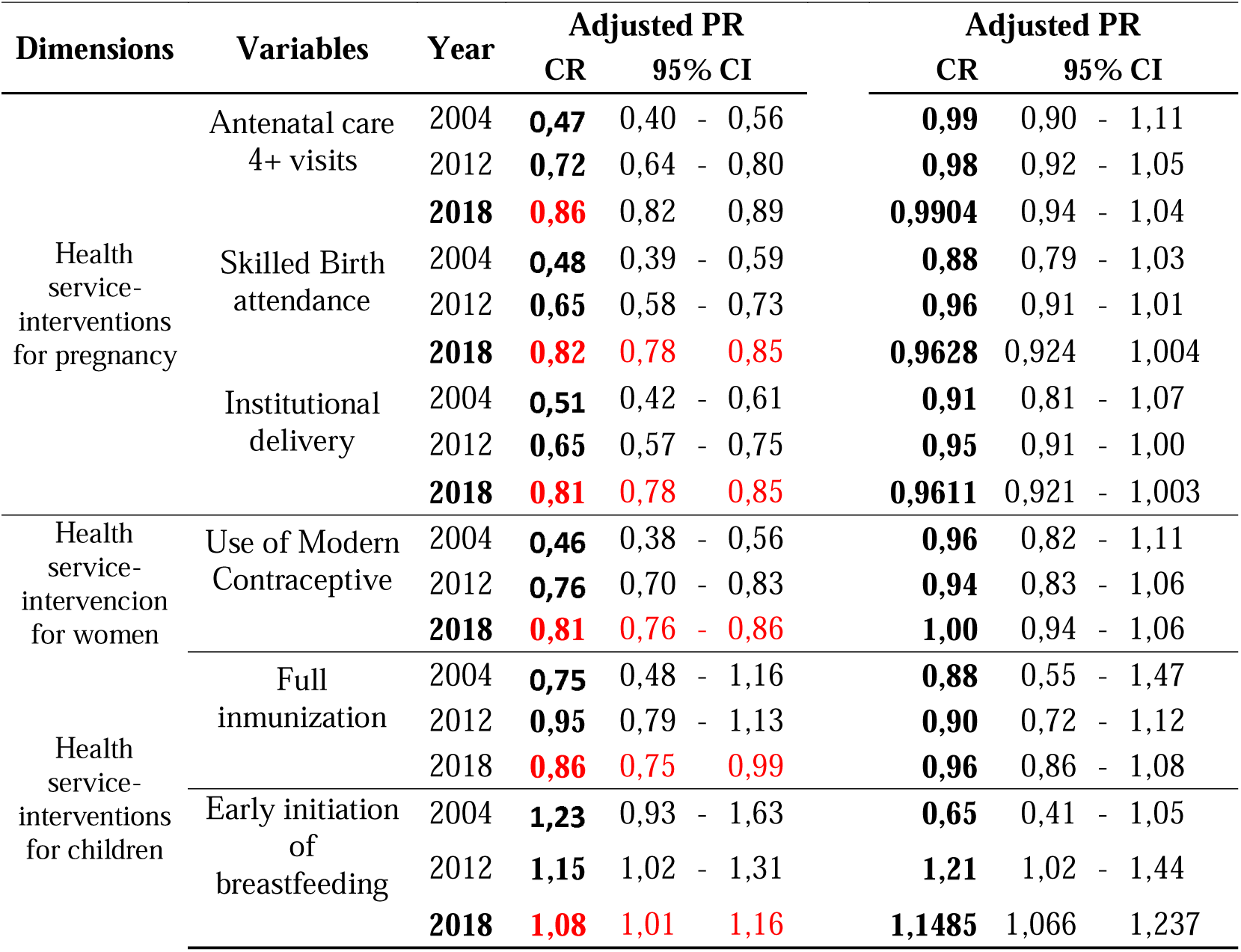
Crude and adjusted coverage rates of six RMNCH interventions in Indigenous and Afro-Ecuadorian women and children compared to the reference group. Ecuador, 2004-2018.

By 2018, the increase in coverage for maternal health services (antenatal care, skilled birth attendance, and institutional delivery) that was observed between 2004 and 2012 continued, albeit at a slightly lower magnitude, around 20 percentage points. However, this trend of increasing coverage did not persist for interventions such as the use of modern contraceptives and full immunization, where increases of 6 and 1 percentage points were observed, respectively. For example, between 2004 and 2012, the use of modern contraceptives rose from 24.3% to 52.9%. A similar situation is observed for these indicators among other ethnic groups.

**Figure 2.**
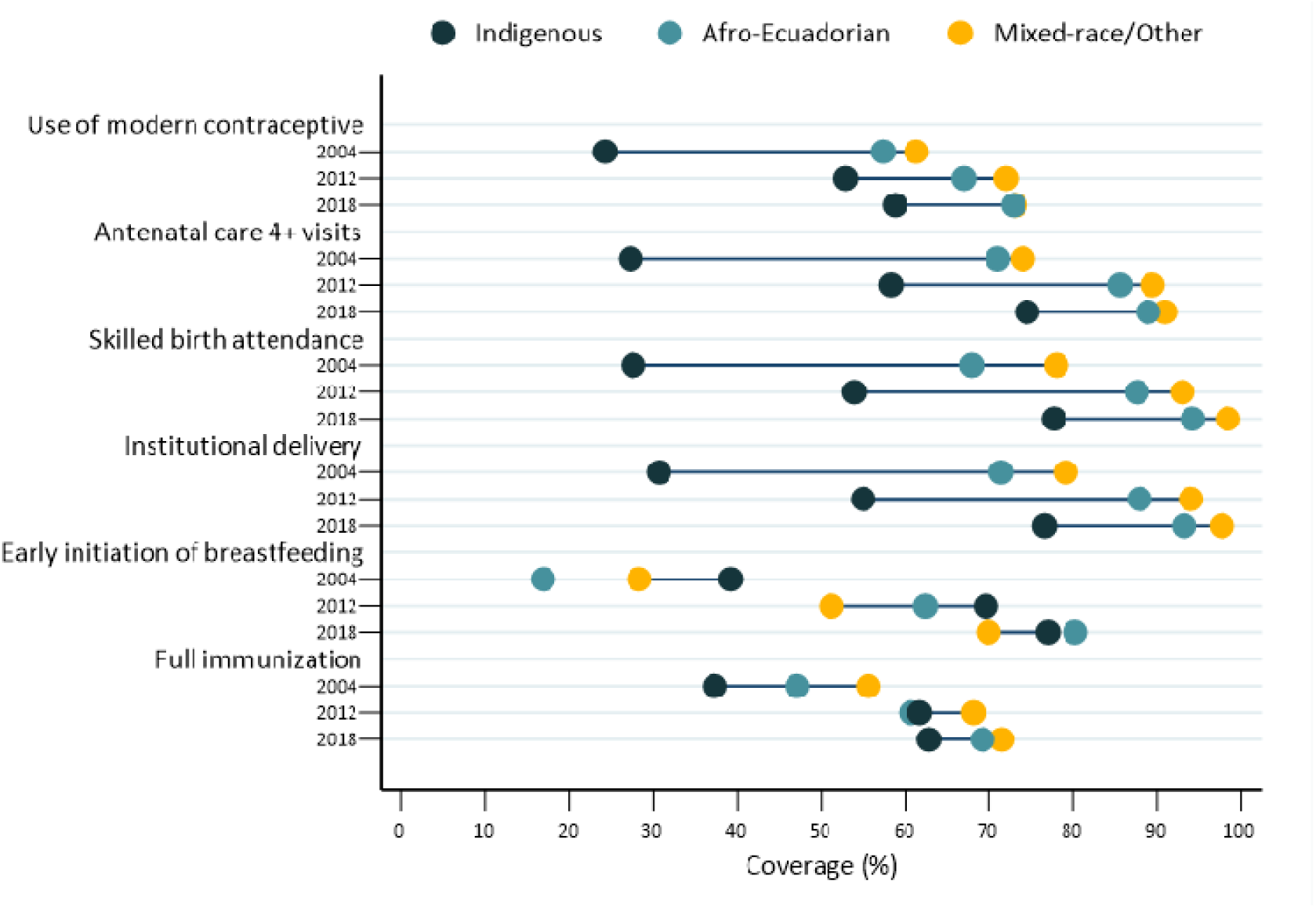
Equiplot coverage and magnitude of inequalities by ethnic groups (2004-2018)

Regarding inequalities, despite notable improvements in coverage between 2012 and 2018, especially for health services, the population self-identifying as Indigenous has been left behind in coverage for almost all interventions from 2004 to 2018. Significant average differences persist, particularly for interventions related to the provision of health services (antenatal care, skilled birth attendance, and institutional delivery, with differences of more than 20 percentage points) and the use of modern contraceptives (14.3 percentage points).

### Results of the Desk Review

Utilizing the predefined keywords, this search yielded 38 articles on Policies and Programs Related to Ethnicity in Ecuador, comprising 11 from PubMed, 8 from Bireme, and 19 manual searches. We found 93 publications on Policies and Programs Related to Maternal and Child Health in Ecuador comprising 31 from PubMed, 34 from Bireme, and 28 manual searches. Following a process of duplicate identification based on author names and titles, the review team removed redundant articles. Subsequently, abstracts were scrutinized applying predefined inclusion and exclusion criteria aligned with the research objectives. 18 articles were selected for inclusion in the study, on Policies and Programs Related to Ethnicity in Ecuador; and 34 articles on Policies and Programs Related to Maternal and Child Health in Ecuador (Details about the search strategy are available in Supplementary Table 4).

**Figure 3:**
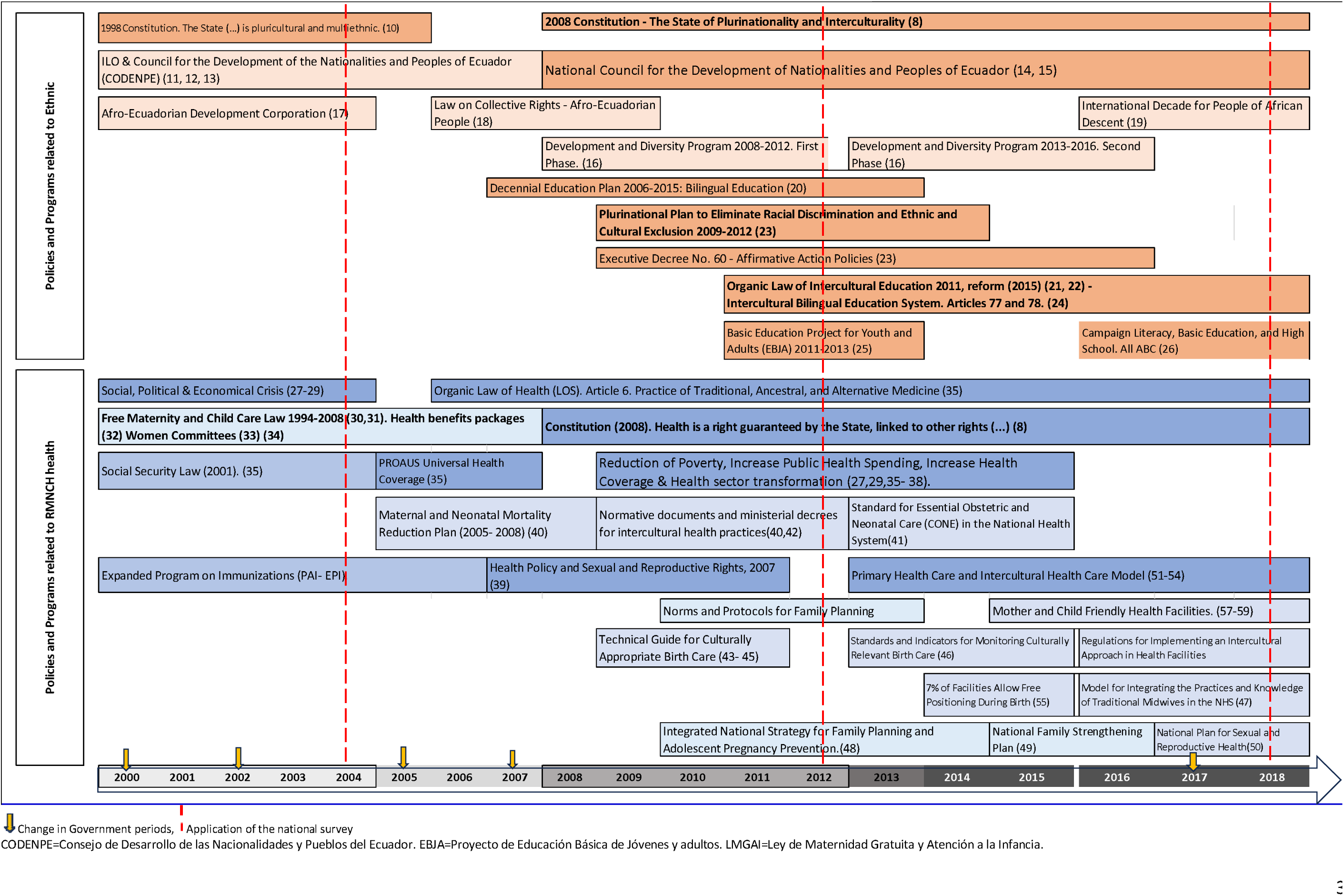
Main policy changes and programmatic activities related to reproductive, maternal, neonatal, and child health in Ecuador, 2004–2018.

### Policies and Programs Related to Ethnicity in Ecuador

Since the 1998 Constitution and as a result of various social mobilizations advocating for the rights of peoples and nationalities, Ecuador was established as a pluricultural and multi-ethnic nation, recognizing the collective rights of its peoples but not their nationalities (10). The 2008 Constitution marked the first time Ecuador was declared a plurinational and intercultural state, making significant advances in inclusive rights for indigenous peoples (8). It established education as an intercultural right (Articles 26 and 27), mandated affirmative action measures to promote equality of rights (Article 11), and guaranteed the collective right to be free from racism and any form of discrimination based on origin, ethnicity, or culture (Article 57). These non-discriminatory educational policies based on culture, skin color, or origin were subsequently incorporated into various normative documents.

The ratification of ILO (International Labour Organization) Convention 169 in Ecuador (Article 2) (11), in 1998 laid the foundation for the establishment of the National Council for the Development of Nationalities and Peoples of Ecuador (CODENPE) (12). CODENPE promoted the sustainable development of Ecuadorian nationalities and peoples through policy formulation, resource allocation, and dialogue with society (13). The National Council for Equality of Peoples and Nationalities (14), created by the Organic Law of National Councils for Equality in 2014 (15), replaced CODENPE and has been active up to the drafting of this study, with additional Development and Diversity Programs 2008–2016 (16). The Afro-Ecuadorian community pursued an independent path by forming the Afro-Ecuadorian Development Corporation in the late 1990s (17). Subsequently, the Law on Collective Rights of the Afro-Ecuadorian People was enacted (18), and the International Decade for People of African Descent was ratified (19).

In the educational sector, the 2006 popular consultation approved eight policies under the Ten-Year Education Plan 2006-2015 (20), which included strategies for inclusive, bilingual education and reducing economic access gaps, such as eliminating tuition fees and providing incentives for uniforms and school materials. The Organic Law of Intercultural Education (LOEI) of 2011 (21), established the state’s contribution at 0.5% annually, aiming to reach at least 6% of GDP (Ten-Year Plan 2006) (22). The “Plurinational Plan to Eliminate Racial Discrimination and Ethnic and Cultural Exclusion (2009-2012),” which began in 2011, included affirmative action policies for equal opportunities, such as quotas (at least 10% of Afro-Ecuadorian, Montubio, and Indigenous students in secondary and higher education) (23), a program to support secondary and higher education (ibid, pp. 45 and 46) (16); and a Bilingual Intercultural Education System (SEIB, Article 77) (24), which helped address historical demands of indigenous peoples for the protection of culture and ancestral languages.

Other identified plans and projects include the Basic Education for Youth and Adults (EBJA) Project 2011-2013 (25), which was completed by 324,894 individuals by 2013, 60% of whom were indigenous people from rural areas of Ecuador who received literacy education in their native language. The “Monseñor Leónidas Proaño Literacy, Basic Education, and High School Campaign: Todos ABC,” implemented in 2017 and continuing to the present (26), has shown a reduction in illiteracy across the country.

### Policies and Programs Related to Maternal and Child Health in Ecuador

The period from 1998 to 2006 in Ecuador was characterized by a deep social, economic, and political crisis, impacting health indicators and coverage of interventions (27–29). Among the social protection policies for maternal and child health during this period, the Free Maternity and Childcare Law (LMGAI) stands out. Essential health benefits packages (30) increased coverage of prenatal care, professional-assisted deliveries, institutional births (30), and access to contraceptive methods (31). This was achieved through financing and implementation mechanisms based on local economic transfers, centralized purchases by international organizations, and strong promotion of social participation, involving indigenous women leaders from rural areas in user committees (32). The Organic Health Law, in effect since 2006 (Art. 6), promotes and enhances the practice of traditional, ancestral, and alternative medicine (33), and in 2006, the PRO-AUS universal health insurance project, but with the change of government, it was not continued (34).

The period from 2006 to 2014 was characterized by economic growth, poverty reduction, and increased public health expenditure, which explains, in part, the increase in RMNCH health intervention coverage (27,35,36) and reduction in maternal and child health disparities (29,37). This was further supported by the health sector transformation process initiated in 2007(27,38), reinforced by the 2008 Constitution, which strengthens the protection of health as a human right (8). Among the main national plans and policies identified are: the “Health, Sexual and Reproductive Rights Policy” (2007) (39) promoted actions against discrimination by ethnicity and recognition, appreciation, and respect for interculturality; in 2008, the “National Plan for the Reduction of Maternal and Neonatal Mortality” (40), and the implementation of the Standard for Essential Obstetric and Neonatal Care (CONE) in the National Health System(41), which coordinated different providers of obstetric and neonatal health services, offering integrated services.

The plurinational nation in the 2008 Constitution promoted the integration of institutional health care with traditional and intercultural practices through normative documents and ministerial decrees for intercultural health practices(40,42). The Technical Guide for Culturally Appropriate Childbirth Care (43) of 2008 emphasized the importance of this adaptation; in 2014, the regulation for the practice of alternative medicine use (44), and in 2017 the Guidelines for the implementation of medicinal gardens in primary care health facilities (45). In 2015, the “Standards and Indicators for Monitoring Low-Risk Normal Birth Care in Free Position in First and Second Level SNS Facilities” was published (46), and a year later, the Manual for Integrating Ancestral Midwifery Practices and Knowledge into the SNS (47). Additionally, a regulation for applying the intercultural approach in SNS health facilities was introduced, and in 2017, guidelines were issued for implementing medicinal gardens in first-level health facilities (45).

Reproductive health plans and strategies have undergone major changes, from the National Intersectoral Strategy and Family Planning and Prevention of Pregnancy in Adolescents (ENIPLA) (48) focused on sexual and reproductive rights, to the so-called “Family Plan” (49); finally, between 2017 - 2021, the National Plan for Sexual Health and Reproductive Health is issued (50).

Between 2014-2018 some reported results were: the incorporation of 1.700 Primary Care Technicians (TAPs) who included indigenous populations (51,52), in the framework of the Comprehensive Health Care Model (MAIS-FCI) (53,54). In 2017, 7% of the first level of care health establishments had birthing rooms with intercultural relevance (55), and 2.460 midwives integrated into the Public Health System (56). As part of the global baby-friendly hospital initiative (57), Ecuador and Brazil are the countries with the most certified establishments between 2008 and 2014 (58); in 2017, the “Health Regulation for Certification as Friends of Mother and Child (ESAMYN)” (59), was issued for SNS establishments that provide childbirth services,

In summary, between 2004 and 2012, the presence of social organizations that advocate for high-level policies on the rights of indigenous peoples is evident. However, the mere existence of these organizations and policies in Ecuador did not reflect sufficient social and health outcomes until several policies and normative documents were added, which should have integrated the intercultural approach into health care within the SNS, within the framework of a Constitution that since 2008 recognizes a plurinational state and reinforces health as a human right. Although some documents with specific results are identified, there are few studies analyzing the impact of the policies or strategies implemented at the national or local level.

## Discussion topics

Few studies analyze health changes over long periods (60,61), and their relationship with changes in government programs or policies. Although a causal relationship between the policies described and the results cannot be established, studies based on other countries have managed to extract important lessons applied to countries in similar contexts(62). Several countries with progressive surveys for decision-making and monitoring interventions have conducted analyses particularly focused on child mortality outcomes (63,64). This is one of the few studies that focuses on coverage of RMNCH health interventions.

Previous studies documented the vast ethnic inequality gaps in maternal and child health intervention coverage in Ecuador in 2004, comparable only to the situation in Guatemala and Peru across all of Latin America (1,2) (65), This situation persisted until 2012, as coverage remained low despite a reduction in inequality gaps. This study shows that between 2012 and 2018, the percentage of RMNCH health intervention coverage tripled (from 25% to 75% between 2004 and 2018). Although the increase is observed across all ethnic subgroups, it is more evident among self-identified indigenous populations. Coverage gaps between groups were progressively reduced, except for immunizations and breastfeeding. Additionally, the percentage of women aged 15 to 49 with secondary education or more increased, and wealth in the highest quintile (Q5) doubled.

These improvements coincide with the generation of inclusive policies, strategies, and affirmative actions, especially from 2008 onwards, supported by the constitutional framework, which may explain the observed improvements in the data over time, similar to other studies, it is observed that economic growth, economic policies and the allocation of funds will also have an impact on the results of proposed health policies(62). In Latin America, only Bolivia and Ecuador have achieved state-level structural reforms following the constitutional declaration of a “plurinational state,” incorporating principles and guidelines that reclaim “Sumaq Kawsay” civilizational models and ways of life. Educational demands emphasized the relevance of basic and higher education (66). Successful strategies that have mitigated health inequalities have addressed socioeconomic disparities and women’s needs by breaking financial barriers (2).

Several programs, plans, and public policies that incorporate intercultural practices in health have been identified. The development and nationwide implementation of protocols, guidelines, and high-level policies (ministerial decrees) are crucial for understanding and training healthcare providers (9). However, few documents have been identified that evaluate the impact of these or normative documents for tracking and monitoring their implementation. Decennial plans and policies take several years to be confirmed as specific laws and programs. It is necessary to monitor coverage of services and quality of care with an equity perspective to assess how different social groups are benefiting from progress in healthcare (67). Routine surveys are helpful for better monitoring and decision-making (4,9).

Key points for improving intervention approaches include involving local actors, traditional midwives (68,69), primary care technicians, and user committees (32). Programs with intercultural facilitators have succeeded when the community is included in the health program process, when relationships are built, such as community networks, programs with traditional elements and cultural approaches, oral knowledge transmission, visual or other easily interpretable communication styles, and pragmatic and collaborative approaches (70). Emphasizing the importance of bringing services closer to communities, strengthening the primary care level, and promoting policy advocacy, community mobilization, and collective action to promote racial equity (71), further encourages women to seek these services (72).

Political stability and sustained economic growth have been identified as facilitating factors in maternal and child health impacts in other countries (9). Increased public spending on health (27,73) as well as improvements in health infrastructure, equipment, and human resources (27,74,75), show positive impacts on health coverage outcomes (35,37,76). These improvements appear to have focused on rural areas, evidenced by the reduction of inequalities in these areas where the highest percentage of the self-identified Indigenous population resides (29,77,78).

Interventions such as exclusive breastfeeding and immunizations require differentiated analysis. The population that identifies as Indigenous is more likely to provide exclusive breastfeeding to their children (79), and the present study shows that this coverage is decreasing, possibly as women move to urban areas. This warrants an analysis of the factors leading to the interruption of breastfeeding in children. The minimal increase in immunization coverage from 2012 to 2018 (only 4 percentage points) may be related to changes in policies and strategies, shifting from the vertical program approach (PAI) to MAIS-FC, which requires further study to better adapt policy changes or national directives (53).

Our findings show that despite advancements to reduce inequalities mainly observed in the first period (2004-2012), they did not maintain the same momentum in the following years. Therefore, by 2018, indigenous populations still have 20% less coverage of maternal and child interventions compared to the reference group. Factors that were not measured in this study persist and explain these gaps (the crude vs. adjusted risk ratios show a significant jump). This is a pre-pandemic snapshot, as progress in coverage was being made, which could have been radically affected by the COVID-19 pandemic.

### Strengths and Limitations

This study has several limitations. As a descriptive ecological correlation study, controlling for all confounding variables was not possible. However, the regression model included key social factors like education, wealth, and rurality. While comparisons were primarily visual, the mixed-methods approach allowed for the analysis of nationally representative data, enabling insights into concurrent policies and generating new hypotheses. Further research is needed to explore key strategies and their impact. Though the reliance on digital evidence may have caused publication bias, efforts were made to reduce this by including grey literature and minimizing strict quality criteria.

### Conclusions

Ecuador is a country with a significant percentage of Indigenous population, which over 14 years has shown a progressive increase in the coverage of RMNCH health interventions and a significant reduction in health disparities in RMNCH, as well as progress in education levels and wealth among mothers who self-identify as Indigenous. Despite these improvements, there are still ethical disparities that need to be addressed.

These improvements align with a period of increased public social investment in health and inclusive education policies, coupled with efforts in social participation and organization that succeeded in incorporating the recognition of plurinationality into the Constitution. This led to the development of strategies to adapt the healthcare system to interculturality, and plans and projects aimed at eliminating racial discrimination and ethnic exclusion, resulting in better education and strengthening of interculturalism. These achievements provide valuable lessons that other countries in the region could benefit from.

The generation of several normative documents in health incorporating an intercultural approach is notable, as well as the involvement of agents or leaders who have contributed to the implementation of strategies at the local level. This underscores the importance of bringing services closer to communities, mobilizing communities, and collective action to promote racial equity.

An important facilitating factor was a period of political stability that coincided with economic growth and investment in social policies in education and health, demonstrating the importance of comprehensive social and economic growth for sustainable health outcomes. Social improvements should now expand their policies with alternatives to strengthen higher education, promote decentralization, and local integrated development to curb urban-rural migration by generating local development alternatives.

Studies with ecological and temporal designs, based on routine surveys, allow for observation of temporal changes in health and contribute to the analysis of the potential impact of implemented strategies. Ongoing monitoring and assessment of the adherence to and impact of programs and policies are required, along with analyzing the economic and health impacts resulting from the economic crisis caused by the COVID-19 pandemic and attempting to reinforce or resume those strategies and initiatives that have proven effective over time.

## Contributor

PR-Q performed the statistical analysis and drafted the entire manuscript. LF contributed to the preparation of the datasets and analyzed the indicators, NA contributed to the preparation of the datasets, statistical analysis, and critical revision of the manuscript and the tables, JE contributed to the search for scientific evidence, improvement of tables and figures. The final manuscript was read and approved by PR-Q, NA, JE, LF. The corresponding author (PR-Q) and NA had full access to all the study data. The corresponding author had final responsibility for the decision to submit for publication

## Funding

This study was not supported by any sponsor or funder. The contribution of the publication was supported by the Central University of Ecuador.

## Declaration of interests

The authors declare that they have no competing interests.

**Technical appendix, statistical code, and dataset available from the Dryad repository,** DOI: https://microdata.worldbank.org/index.php/catalog/979 to RHS 2004. DOI: https://www.ecuadorencifras.gob.ec/encuesta-nacional-de-salud-salud-reproductiva-y-nutricion-ensanut-2012; to ENSANUT 2012, and DOI: https://www.ecuadorencifras.gob.ec/salud-salud-reproductiva-y-nutricion/, to ENSANUT, 2018.

## Supporting information

https://1drv.ms/w/s!AoE-dzPQT9eWisACla9xfdbMcECqzg?e=WIQxwr

## Data Availability

https://microdata.worldbank.org/index.php/catalog/979
https://www.ecuadorencifras.gob.ec/encuesta-nacional-de-salud-salud-reproductiva-y-nutricion-ensanut-2012
https://www.ecuadorencifras.gob.ec/salud-salud-reproductiva-y-nutricion/

https://www.ecuadorencifras.gob.ec/encuesta-nacional-de-salud-salud-reproductiva-y-nutricion-ensanut-2012

https://www.ecuadorencifras.gob.ec/salud-salud-reproductiva-y-nutricion/

## Notes

### Competing Interest Statement

The authors have declared no competing interest.

### Author Declarations

https://microdata.worldbank.org/index.php/catalog/979 https://www.ecuadorencifras.gob.ec/encuesta-nacional-de-salud-salud-reproductiva-y-nutricion-ensanut-2012 https://www.ecuadorencifras.gob.ec/salud-salud-reproductiva-y-nutricion/

